# Concurrent malaria and dengue fever in (sub-Saharan) Africa: a systematic review and meta-analysis

**DOI:** 10.1101/2022.05.24.22275526

**Authors:** Tewelde T. Gebremariam, Zeleke Mekonnen, Jonas B. Danquah

## Abstract

**Objectives:** To determine the prevalence of concurrent malaria and dengue fever, aetiologies, and the association between the two infections in Africa.

**Methods:** We assessed evidence from previous studies in Africa that were available in MEDLINE and EMBASE databases between January 01, 2005, and March 30, 2022.

**Results:** A total of 3942 studies were identified from the databases of which 30 studies from 13 African countries, including 25,241 febrile patients, were included in the meta-analysis. The result of the meta□analysis showed a 4.0% pooled prevalence of concurrent malaria and dengue fever in (sub-Saharan) Africa, and the highest rate was recorded in Central Africa (5.8%) followed by East Africa (4.7%) and West Africa (2.8%). Children <18 years old (57%) and females (52.8%) were more affected by the co-infection than adults and females, respectively. The dengue virus serotypes identified were DENV-2, DENV-3, DENV-1, and DENV-4 in descending order with a proportion of 39%, 31%, 27%, and 3%, respectively. Moreover, *Plasmodium falciparum* was the only specified malaria parasite in the co-infection among the included studies. Significantly higher odds of malaria infection were documented due to dengue fever when compared with malaria mono-infection. However, no significant odds of acute dengue co-infection were recorded due to malaria in contrast to dengue mono-infection.

**Conclusion:** This study showed a high prevalence of concurrent malaria and dengue fever in Africa. Healthcare workers should bear in mind the possibility of dengue infection as differential diagnoses for acute febrile illness as well as the possibility of co-existent malaria and dengue in endemic areas. Also, high-quality multi-centre studies are required to verify the above conclusions.

Protocol registration number: <u>CRD42022311301</u>.

## INTRODUCTION

Acute undifferentiated febrile illness (AUFI) is one of the most frequent reasons for healthcare seeking in Africa.^1^ AUFI usually begins with non-specific symptoms such as the sudden onset of fever which fewer progresses to prolonged duration, headache, chills, and myalgia which may later involve specific organs. It can range from mild and self-limiting illness to advancing, deadly disease.^2^ Malaria and dengue fever are leading causes of AUFI.^3^

Africa carries the highest global malaria burden, with 2000 million cases (92%) in 2017 alone.^4^ Human malaria is mainly caused by four *Plasmodium* species, namely *P. falciparum, P. vivax, P. malariae*, and *P. ovale* with variable geographic distribution. *P. falciparum* accounts for nearly all malaria deaths in sub-Saharan Africa, which bears over 90% of the global malaria burden.^5^ Likewise, the prevalence of dengue in the region has dramatically increased over the past few decades although this specific infection is neither systematically investigated nor generally considered by clinicians.^6^ In 2013, about 16 million apparent and over 48 million inapparent dengue were estimated to have occurred, and most countries in the continent reported recurrent outbreaks.^7^ Similarly, dengue fever is caused by four genetically distinct dengue viruses (serotypes 1-4).^8^

Though malaria or dengue virus mono-infection can be severe, concomitant infections could be even more fatal.^9, 10^ The two mosquito-borne diseases have an overlapping epidemic pattern in Africa.^11^ Due to their similar clinical presentation, possible concurrent malaria-dengue fever is often neglected ^12^ and generally misdiagnosed as malaria only.^6, 13^ Misdiagnosis is more probable during co-infection than mono-infection and this may result in slow identification of dengue fever outbreaks with potentially high morbidity and mortality.^6, 14^

This review aimed to gather evidence to answer the question: how common is *Plasmodium* and dengue virus co-infection in Africa could be? The specific review objectives were (1) to determine the prevalence of concurrent malaria and acute dengue in Africa (by region and demographic characteristics), and (2) to describe the dengue virus serotypes in the concurrent infection.

## METHODS

The protocol of the review was registered in the International Prospective Register of Systematic Reviews, PROSPERO (CRD42022311301) and followed the Joanna Briggs Institute (JBI)^15^ and the Preferred Reporting Items for Systematic Reviews and Meta-Analyses (PRISMA)^16^ checklists (see online supplementary research checklists).

### Inclusion and exclusion criteria

Case report, cross-sectional and case-control studies that reported concurrent malaria and dengue fever among uncomplicated febrile cases in African regions were included. According to the United Nations, Africa is divided into five regions: Northern Africa, Central or Middle Africa, Southern Africa, East Africa, and Western Africa.^17^ Similarly, the World Bank lists a total of 48 countries in the sub-Saharan African region.^18^

Malaria might be diagnosed by malaria rapid diagnostic tests or microscopy and/or polymerase chain reaction while dengue fever might be identified through antigen/antibody test and/or reverse transcriptase-polymerase chain reaction. Acute dengue or dengue fever was defined as positive for dengue IgM or NS1 antigen test or RT-PCR.

Articles of which full texts could not be obtained or were not available in English were excluded.

The primary outcome measure was the prevalence of concurrent malaria and dengue infection in Africa. The secondary outcomes were: (1) the proportion of males and females with concurrent malaria and acute dengue, and (2) the proportion of dengue virus serotypes in the co-infection. The literature screening and selection process is depicted in Figure 1.

**Figure 1.**
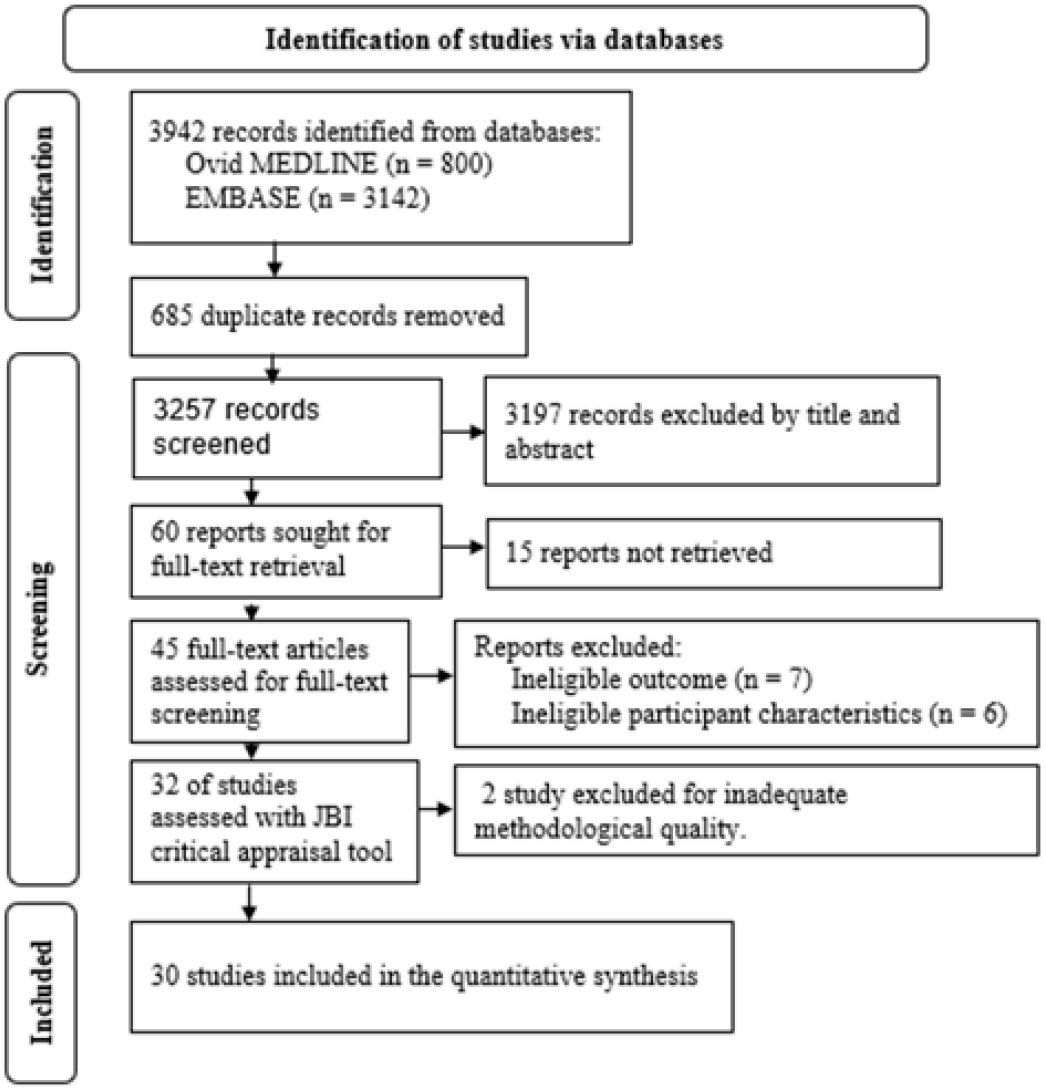
Flowchart showing the identification and selection process of articles for inclusion in the review.

### Databases and search strategy

The CoCoPop mnemonic (condition, context, and population)^19^ was used to formulate the review question and systematically search all relevant studies published between January 01, 2005, to March 30, 2022, from MEDLINE and EMBASE databases. The search strategy used was: **(exp Malaria, Falciparum/ or Malaria/ or ((*Plasmodium falciparum* or *Plasmodium* infect* or malignant tertian or (malaria or marsh or remittent)) adj3 fever).ti**,**ab.) and (exp Dengue/ or exp Dengue Virus/ or ((dengue or Break?bone) adj3 fever).ti**,**ab.)**.

### Study quality appraisal and data extraction

The Joanna Briggs Institute System for the Unified Management, Assessment and Review of Information (JBI SUMARI) tool^20^ was used to screen each article and extract relevant data for the review. Two of the authors (TT, JD) independently screened each article at an abstract and full-text level. The discrepancy between the two reviewers was resolved through discussion. Articles endorsed in the full-text screening were subjected to JBI critical appraisal tool. Those with good quality scores were undergone data extraction. Data extraction included the first author’s last name, publication year, country/region of study, sample size, number of concurrent malaria and dengue infections, demographic characteristics (age, gender) of patients with concurrent infections, and dengue serotypes in the concurrent infections. The JBI criteria were used to score the quality of each study. Studies with a score greater than or equal to four were seen as good quality to be included in the meta-analysis.

### Statistical analysis

The meta-analysis was performed using Jamovi software (version 2.2.5)^21^, OpenMeta-analyst^22^, and the JBI SUMARI tool.^20^ The random-effect model (DerSimonian-Laid) and the effect size model measures of raw proportion/Freeman-Tukey double arcsine transformed proportion were used to determine the prevalence/proportion estimates. The heterogeneity of included studies was measured using Tau, Tau^2^, I^2^ statistic, and Cochran’s Q-test. Forest plot was used to visually assess the prevalence and odds ratio with their 95% confidence intervals (CIs). Publication bias was evaluated using Egger’s regression test and Rank Correlation Test. P-value <0.05 was considered statistically significant.

### Patient and public involvement

This study was done without patient or public involvement.

## RESULTS

### Literature retrieval and article characteristics

A total of 3942 records were identified during literature retrieval from the databases. A total of 30 studies from (sub-Saharan) Africa involving 25,241 febrile patients were included in this meta-analysis ^23-53^ (Figure 1, Table 1). The quality score of the included studies ranged from 4 to 7 on a maximum scale of quality scores of 8 for case reports, 9 for cross-sectional studies, and 11 for case-control studies using the JBI criteria (Table 1).

**Table 1.**
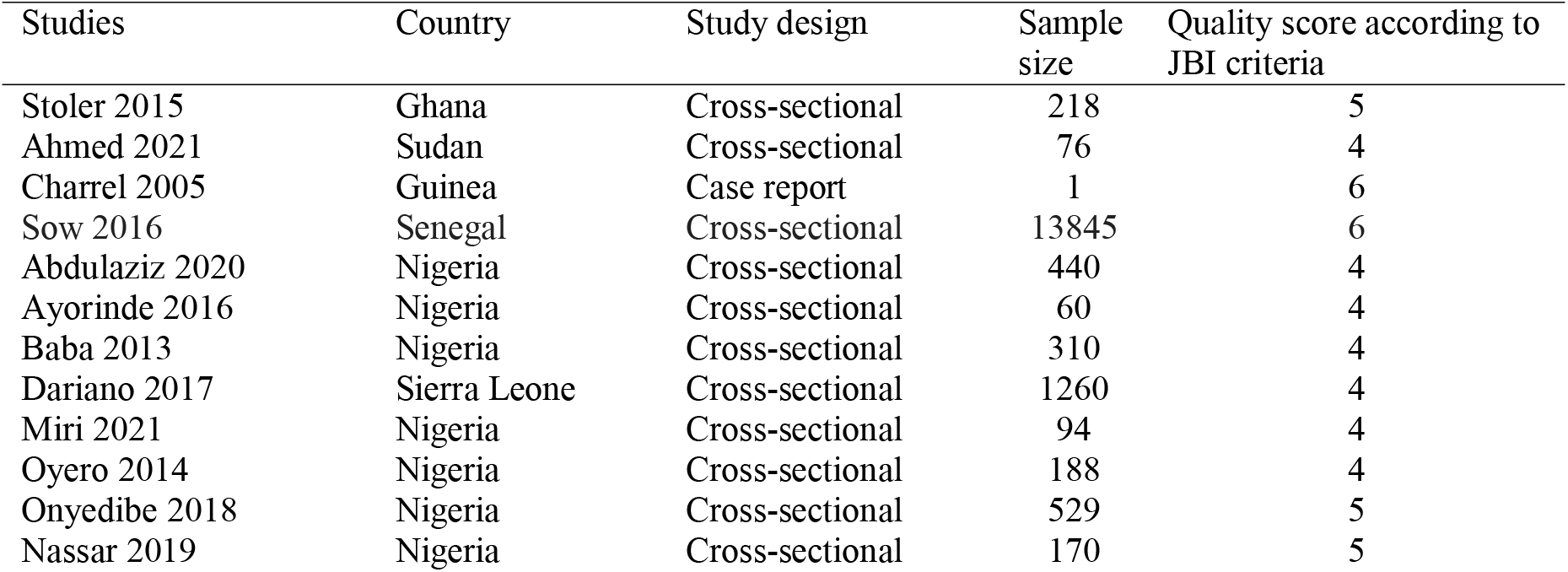
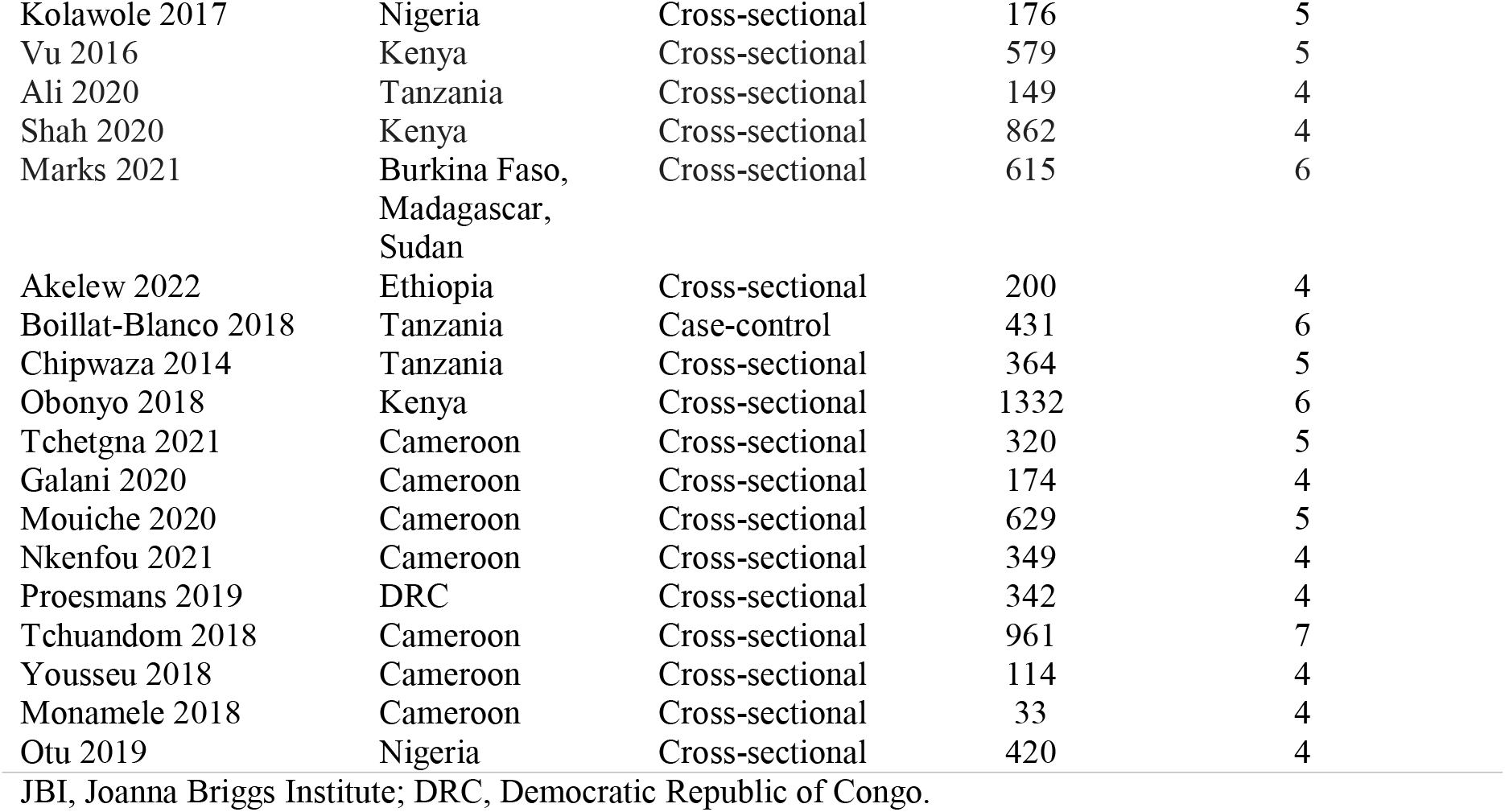
Basic Characteristics and quality score of included studies.

### Concurrent falciparum malaria and dengue fever

The random-effect (DerSimonian-Laird) model estimator and raw proportion and/or Freeman-Tukey double arcsine transformed proportion were used in the meta-analysis. The prevalence of malaria and dengue co-infection in (sub-Saharan) Africa per 1000 febrile cases was 40 (95% CI, 32 – 48, SE = 0.004) with significant heterogeneity (Tau= 0.018, Tau^2^ = 0.00, I^2^ = 95.52%, and Q = 625.28, p < 0.001) (Figure 2).

**Figure 2.**
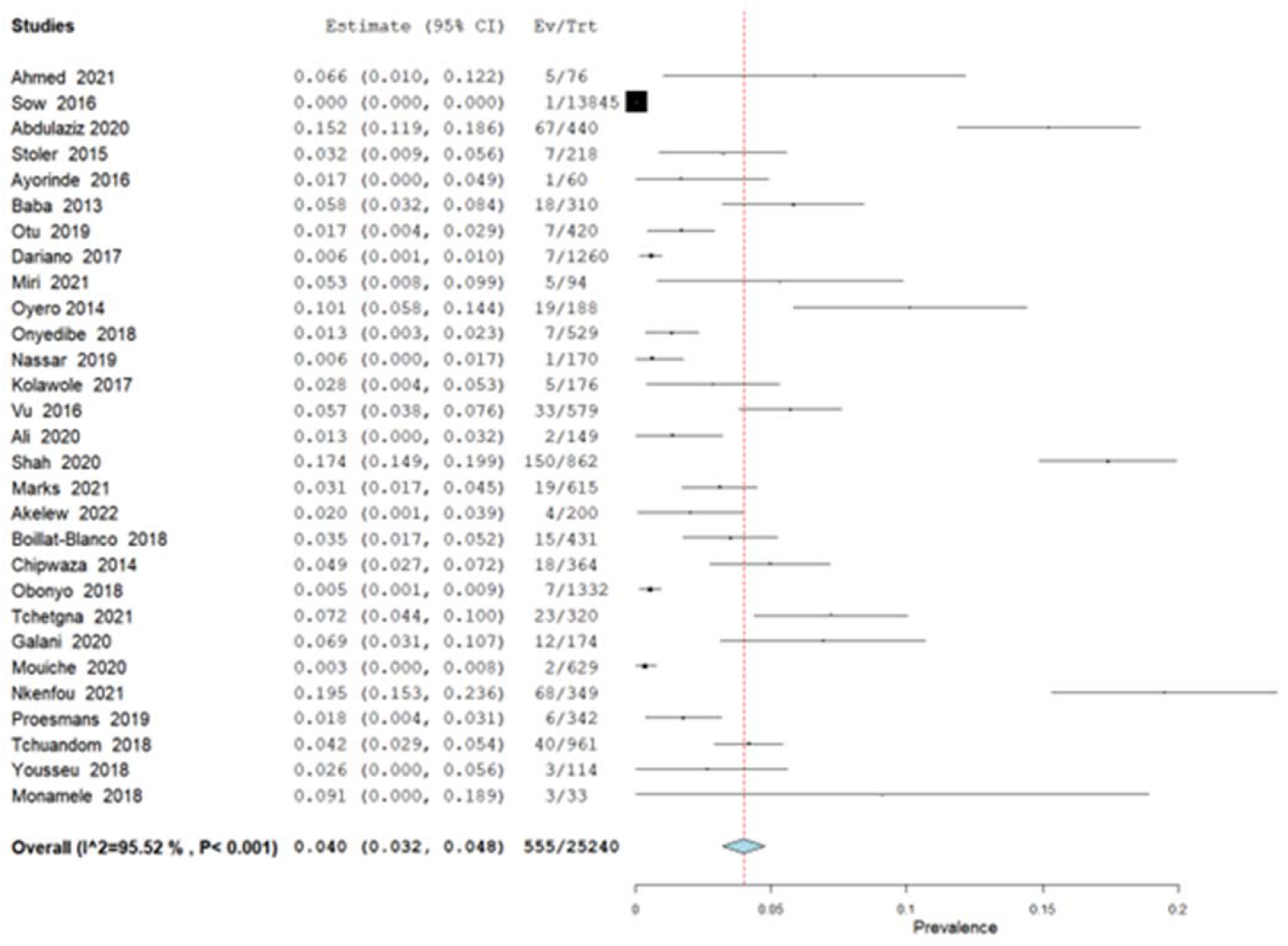
Forest plot delineating prevalence of malaria and acute dengue co-infection in (sub-Saharan) Africa.

The prevalence significantly varies across the three African regions (χ^2^ = 537.13, p < 0.00001 and ranges from 28.5 per 1000 febrile cases (95% CI, 18 – 39, SE = 0.0052) in West Africa (Figure 3), 47 (95% CI, 19 – 75, SE = .0142) in East Africa (Figure 4) to 58 (95% CI, 30 – 86, SE = 0.0143) in Central Africa (Figure 5) (Table 2). A case report^23^ was excluded from the prevalence estimation.

**Figure 3.**
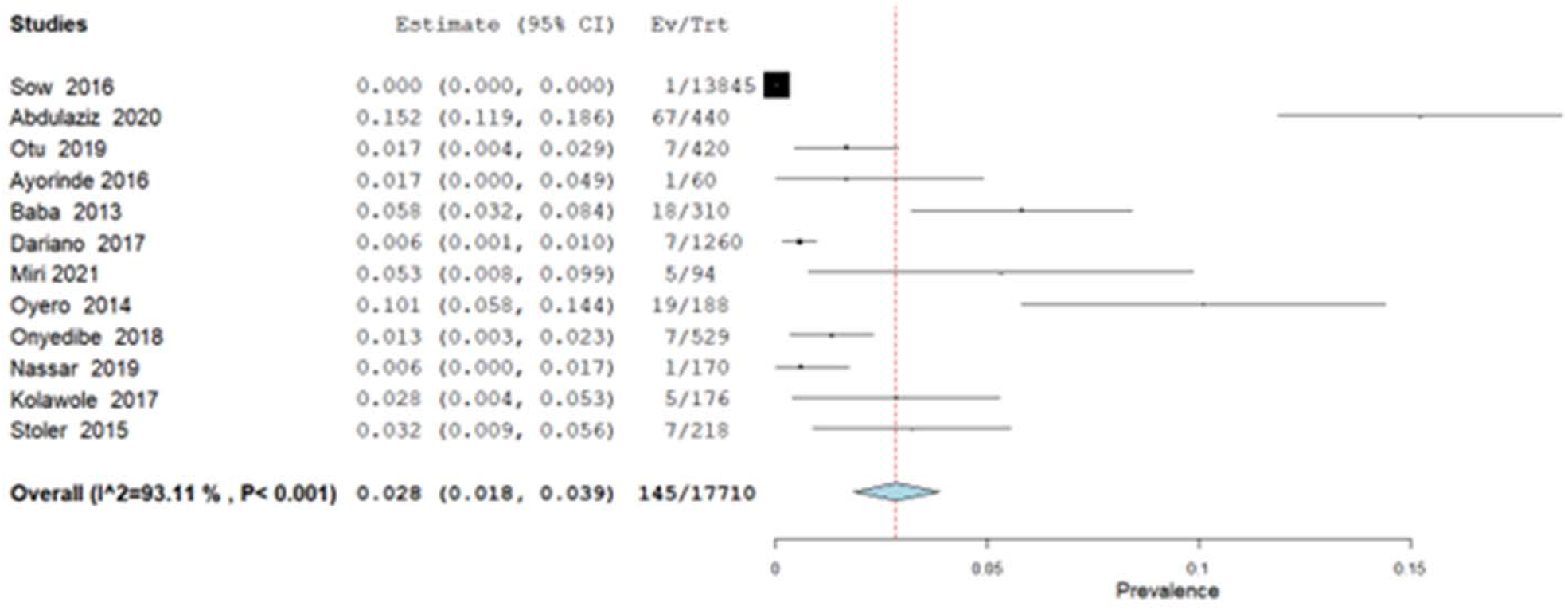
Forest plot showing the prevalence of malaria and acute dengue co-infection in West Africa.

**Figure 4.**
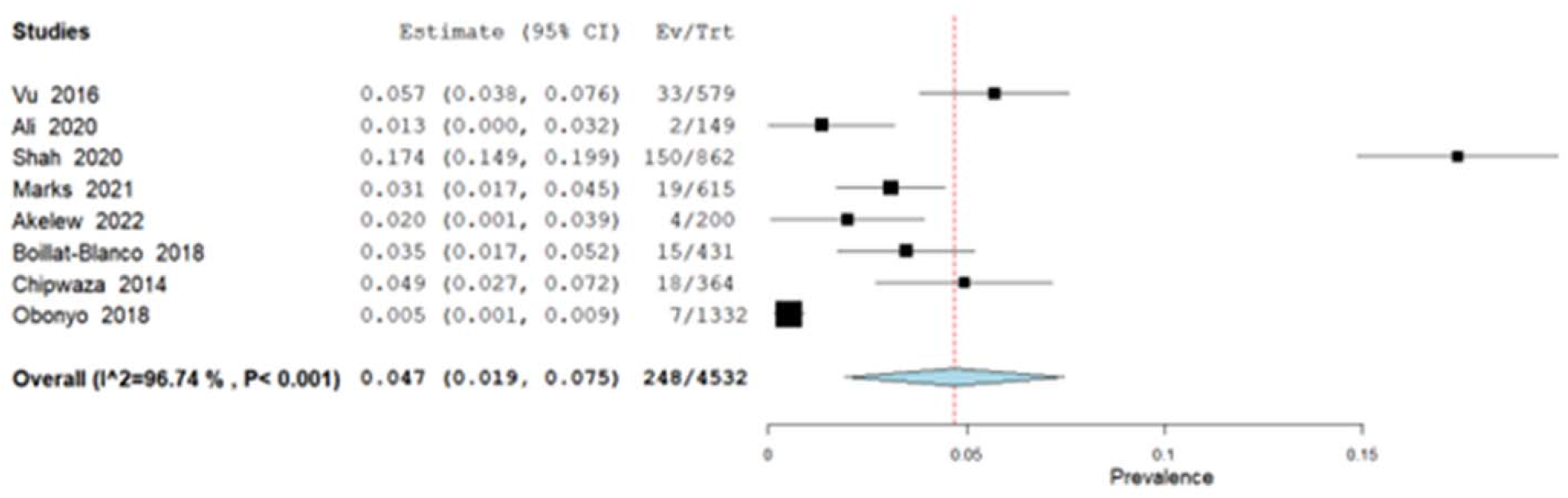
Prevalence of malaria and dengue co-infection in East Africa.

**Figure 5.**
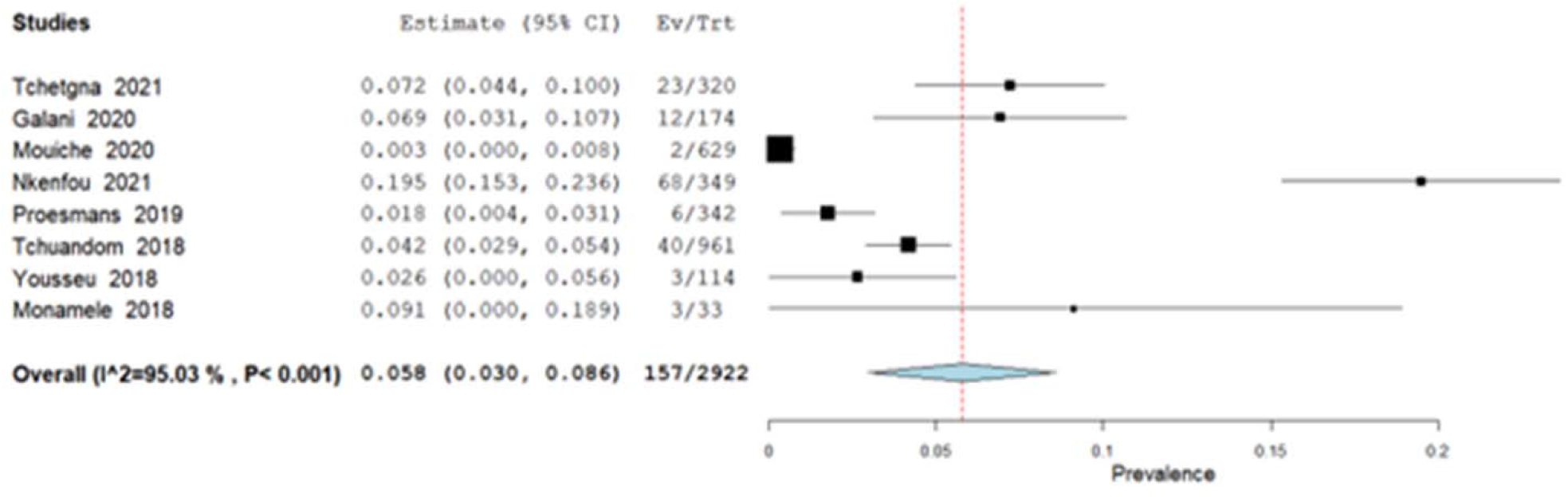
Forest plot depicting prevalence of concurrent malaria and acute dengue in Central Africa.

**Table 2.**
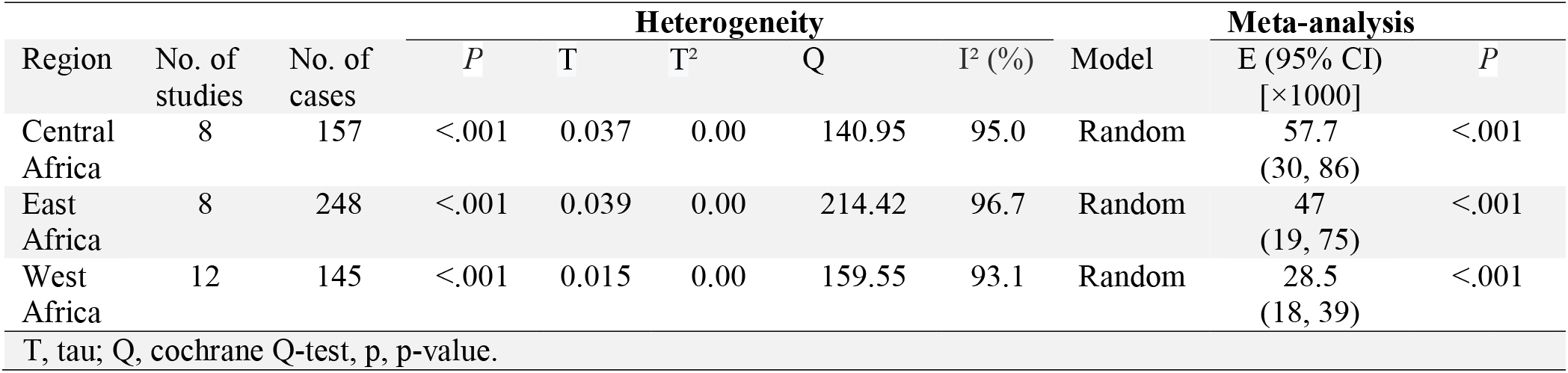
Meta-analysis of concurrent malaria and acute dengue prevalence among febrile patients in three African regions.

Twenty-two studies examined the association between acute dengue virus infection with malaria. The peto’s odds ratio^54^ was used to pool the odds ratio of individual studies. In our meta-analysis (Figure 6), cross-sectional studies in Cameroon^38, 53^, and Kenya^30^ revealed significantly lower odds of co-infection in contrast with the absence of acute dengue infections (OR: 0.60; 95% CI, 0.42 – 0.87). Cross-sectional studies in Nigeria^24, 26, 28, 33, 40, 41, 43^, Tanzania^25^, Sierra Leone^31^, Democratic Republic of Congo (DRC)^42^, Cameroon^39^, Sudan^48, 50^, and Senegal^29^ indicated no significant odds of co-infection in comparison with acute dengue negatives (OR = 1.07; 95% CI = 0.58, 1.94). Studies in Ethiopia^52^, Nigeria^37^, Tanzania^44^, Cameroon^51^, and Kenya^47^, on the other hand, indicated significantly higher odds of concurrent infection to acute dengue negatives (OR = 4.56; 95% CI = 3.29, 6.33). There was no indication of a potential outlier (studentized residuals >±3.0654) and overlay influence (cook’s distance) among the studies. Above all, the summary estimate derived from 22 studies showed significantly higher odds of malaria infection among patients in the absence of acute dengue than acute dengue virus co-infection (OR = 1.51; 95% CI = 1.31, 1.75) with significant heterogeneity (I^2^ = 89; p < 0.001). The 95% prediction interval for the true outcomes (log peto’s odds ratio) was -1.41 to 2.46.

**Figure 6.**
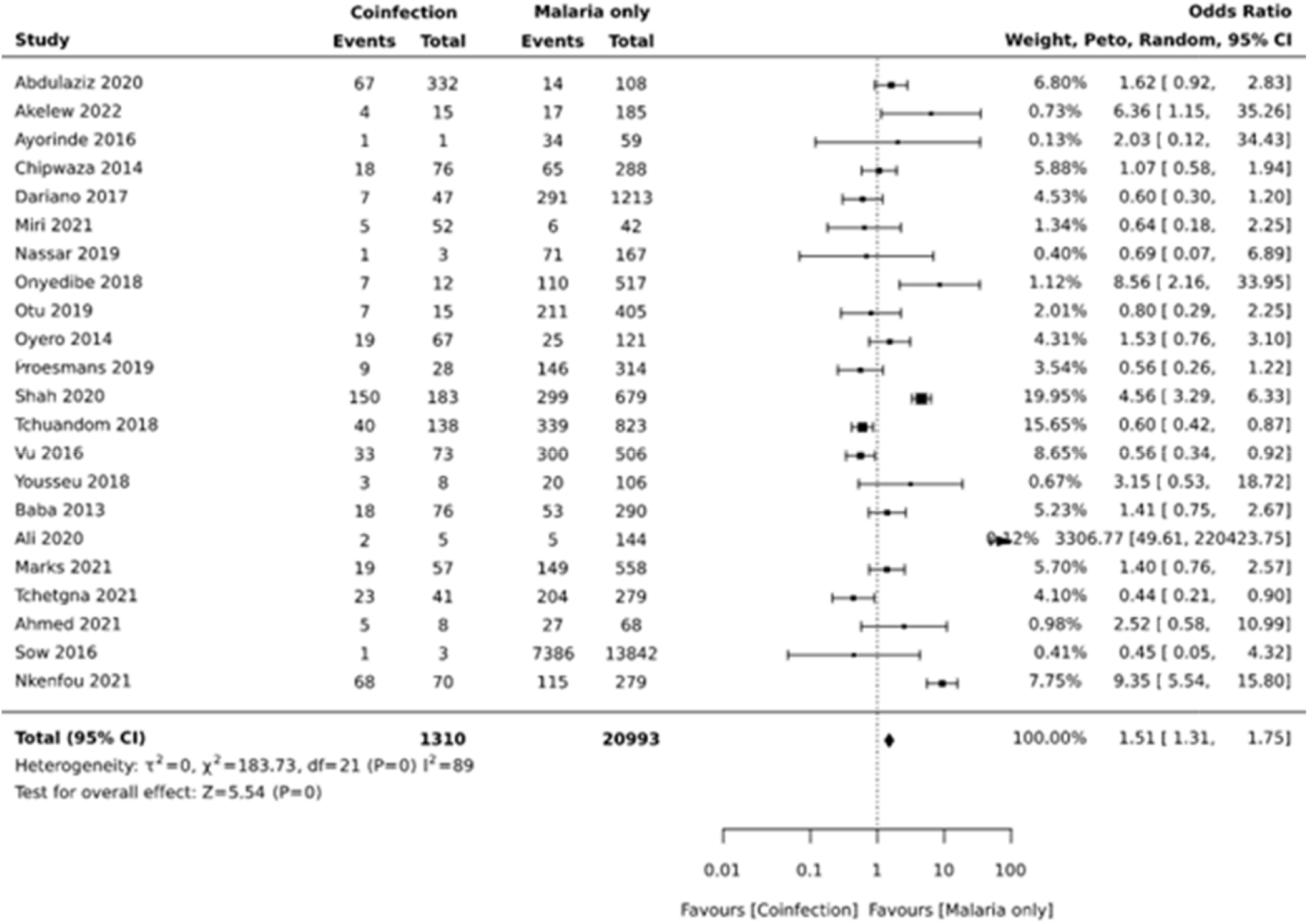
Forest plot: the difference in the prevalence of malaria due to acute dengue virus infection and malaria mono-infection.

Similarly, the odds of acute dengue virus infection were also compared with the presence and absence of malaria infection (Figure 7). Cross-sectional studies in Kenya^30, 47^ and Cameroon^38, 53^ reported significantly low odds of acute dengue in the presence of malaria than in its absence (OR = 0.57, 95% CI, 0.40 – 0.82). Similar studies Nigeria^26, 28, 33, 40, 41, 43^, Tanzania^25^, Sierra Leone^31^, DRC^42^, Cameroon^39^, Sudan^48, 50^, and Senegal^29^; however, showed no notable odds of acute dengue infection in the presence of malaria when compared to the absence of malaria (OR = 1.07, 95% CI, 0.58 −1.94). On the contrary, cross-sectional studies in Ethiopia^52^, Nigeria^24, 37^, Tanzania^44^, and Cameroon^51^ indicated higher odds of acute dengue infection in the presence of malaria than in its absence (OR = 6.98, 95% CI, 2.89 – 16.85). The overall estimate based on 22 studies showed no significant odds of acute dengue co-infection in contrast to malaria mono-infection (OR = 1.00, 95% CI, 0.87 – 1.16) with significant heterogeneity (I^2^ = 86, P <0.0001). The 95% prediction interval for the true outcomes (log peto’s odds ratio) was -1.45 – 2.127.

**Figure 7.**
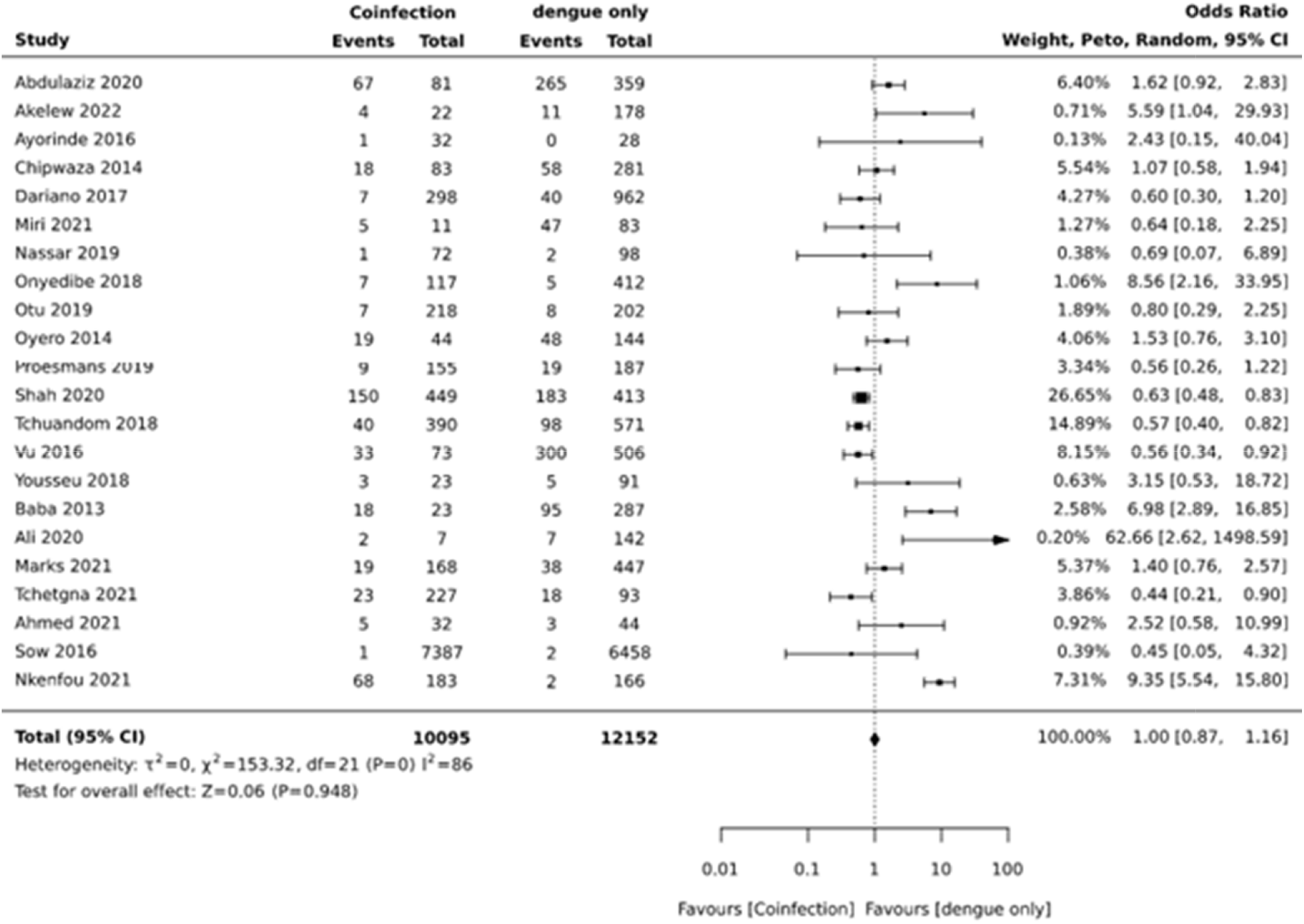
Forest plot showing the difference in the prevalence of acute dengue co-infection due to malaria and acute dengue mono-infection.

The gender distribution among malaria and acute dengue virus co-infection showed that the proportion of females was 52.8% (95% CI, 44.1 – 61.5, SE = 0.044) without evidence of heterogeneity (I^2^ = 0%, p = 0.62), while the proportion of males was 47.2% (95% CI, 38.5 – 55.9, SE = 0.044) with no evidence of heterogeneity (I^2^ = 0%, p = 0.62) (Table 3). Furthermore, the proportion age category of the coinfection was 57.0% (40.9-73.1, SE = 0.082) among children <18 years old and 40.9% (95% CI, 24.5 – 57.0, SE = 0.082) in adults with significant heterogeneity (p <0.001).

**Table 3.**
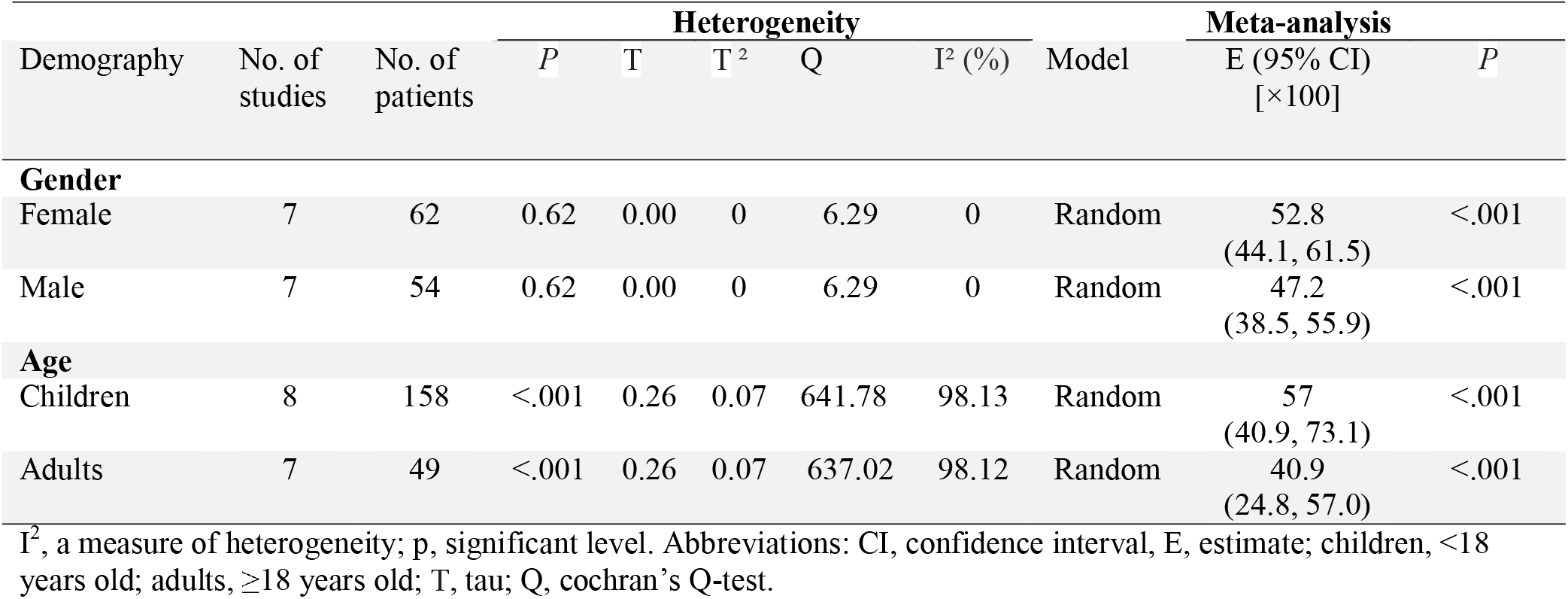
Meta-analysis of demographic characteristics of patients with concurrent malaria and dengue virus infection in (sub-Saharan) Africa.

| Demography | No. of studies | No. of patients | *P* | T | T<sup>2</sup> | Q | I<sup>2</sup> (%) | Model | E (95% CI) [×100] | *P* |
| --- | --- | --- | --- | --- | --- | --- | --- | --- | --- | --- |
| **Gender** |  |  |  |  |  |  |  |  |  |  |
| Female | 7 | 62 | 0.62 | 0.00 | 0 | 6.29 | 0 | Random | 52.8<br>(44.1, 61.5) | <.001 |
| Male | 7 | 54 | 0.62 | 0.00 | 0 | 6.29 | 0 | Random | 47.2<br>(38.5, 55.9) | <.001 |
| **Age** |  |  |  |  |  |  |  |  |  |  |
| Children | 8 | 158 | <.001 | 0.26 | 0.07 | 641.78 | 98.13 | Random | 57<br>(40.9, 73.1) | <.001 |
| Adults | 7 | 49 | <.001 | 0.26 | 0.07 | 637.02 | 98.12 | Random | 40.9<br>(24.8, 57.0) | <.001 |

I<sup>2</sup>, a measure of heterogeneity; p, significant level. Abbreviations: CI, confidence interval, E, estimate; children, <18 years old; adults, ≥18 years old; T, tau; Q, cochrane's Q-test.

The meta-analysis on the proportion of aetiologies of concomitant malaria and acute dengue is shown in Table 4. All the four dengue virus serotypes were identified, where DENV-2, DENV-3, DENV-1, and DENV-4 were documented in descending order with a proportion of 39.5% (95% CI, 16.8-62.2, SE = 0.116), 31.6% (95% CI, 16.0 – 47.2, SE = 0.080), 27.3% (95% CI, 14.6 – 40.1, SE = 0.065), and 3.7% (95% CI, 1.0 – 6.3, SE = 0.014), respectively. The aetiology of malaria among the co-infection was specified in 50.7% (95% CI, 29.6 – 71.9, SE = 0.108) of the patients, and all were due to *P. falciparum*.

**Table 4.** Meta-analysis of the proportion of aetiologies of concomitant malaria and acute dengue in (sub-Saharan) Africa.

|  |  |  | Heterogeneity |  |  |  |  | Model | Meta-analysis |  |
| --- | --- | --- | --- | --- | --- | --- | --- | --- | --- | --- |
|  | No. of studies | No. of patients | *P* | T | T<sup>2</sup> | Q | I<sup>2</sup> (%) |  | E (95% CI)<br>[×100] | *P* |
| DENV serotypes |  |  |  |  |  |  |  |  |  |  |
| DENV-2 | 7 | 97 | <.001 | .39 | .15 | 331.3 | 96.4 | Random | 39.5<br>(16.8, 62.2) | <.001 |
| DENV-3 | 7 | 36 | <.001 | .25 | .06 | 216.2 | 94.5 | Random | 31.6<br>(16.0, 47.2) | <.001 |
| DENV-1 | 6 | 33 | <.001 | .20 | .04 | 118.1 | 89.8 | Random | 27.3<br>(14.6, 40.1) | <.001 |
| DENV-4 | 3 | 7 | 0.81 | .00 | 0 | 7.7 | 0 | Random | 3.7<br>(1.0, 6.3) | .01 |
| Plasmodium species |  |  |  |  |  |  |  |  |  |  |
| Pf | 16 | 305 | <.001 | .57 | .33 | 15549.5 | 99.8 | Random | 50.7<br>(29.6, 71.9) | <.001 |
| NS | 14 | 251 | <.001 | .58 | .33 | 15549.5 | 99.8 | Random | 49.3<br>(28.1, 70.4) | <.001 |

I<sup>2</sup>, a measure of heterogeneity; p, significant level. Abbreviations: CI, confidence interval; E, estimate; Pf, *Plasmodium*

### Publication bias

The publication bias of included studies was evaluated using the funnel plot asymmetry tests: the Egger’s regression test and rank correlation test based on the Freeman-Tukey double arcsine transformed proportion effect size model measures for prevalence/proportion estimates and log odds ratio peto’s method for the difference in the prevalence rates. Both tests showed nonsignificant publication bias (p >0.05) among the studies included in all the analyses.

## DISCUSSION

The first report of malaria and dengue virus co-infection in Africa was documented in 2005.^23^ Our meta-analysis included 25,241 febrile patients from 30 studies conducted in 13 African countries (Ghana, Sudan, Senegal, Nigeria, Guinea, Sierra Leone, Kenya, Tanzania, Ethiopia, Cameroon, DRC, Burkina Faso, and Madagascar) for over around 17 years. As far as we know, this is the first description and analysis of the prevalence, demographic characteristics, and dengue virus serotype profiles of concurrent malaria and dengue infection in (sub-Saharan) Africa.

Based on our analysis, the pooled prevalence of concurrent malaria and dengue was 4.0%, and the highest rate was recorded in Central Africa (5.8%), followed by East Africa (4.7%) and West Africa (2.8%). This result is lower than the finding of a study^55^ on a meta-analysis of severe malaria and dengue co-infection which estimated a prevalence of 32%. The variation could be due to the differences in the study population, model estimator employed, and/or geography of the primary studies included, where our analysis was focused on studies from Africa while the other study included studies from all over the globe. Besides, uncomplicated febrile cases were included in our analysis, unlike above the study which estimated severe malaria prevalence among the co-infection.

In our study, women and children were more affected groups by the concomitant infection than males and adults, respectively. Women and children are more susceptible to mosquito-borne illnesses because they are exposed to mosquito bites for longer periods during dangerous hours ^56^. Moreover, malaria^57^ and dengue^58^ mainly affect children due to underdeveloped specific immunity to infection.^57^

In our analysis, the most prevalent dengue virus serotypes present in the concurrent infection were serotypes 2 and 3. The DENV-2 serotype is divided into six genotypes: Asian I, Asian II, American, Asian/American, Cosmopolitan, and Sylvatic.^59^ The cosmopolitan genotype of DENV-2 is the most widely distributed dengue virus worldwide. The DENV-3 serotype of genotype III is found in Africa.^60^ Furthermore, *Plasmodium falciparum* was the only malaria parasite specified in the coinfection among the included studies. It is known that nearly all malaria cases in (sub-Saharan) Africa are caused by *P. falciparum*.^5^

Our study also reported significantly higher odds of malaria infection among patients with malaria mono-infection than acute dengue virus co-infection. This could be due to the inter-species cross-protection of dengue virus infection against malaria.^11^ Our finding does not corroborate the reports of Kotepui et al. 2019^55^ and Kotepui et al. 2020^61^. Kotepui et al. 2019 showed significantly lower odds of co-infection while Kotepui et al. 2020 reported no notable difference in co-infection due to acute dengue compared to malaria mono-infection. Our study further elucidated no significant odds of acute dengue virus co-infection infection due to malaria than acute dengue virus mono-infection. Our report is in disagreement with the findings of Kotepui et al. 2020^61^ which indicated higher odds of acute dengue due to malaria co-infection in contrast to dengue virus mono-infection. The discordant findings could be attributed to the variation in model estimator/effect size model measures and/or quality/geography of included studies.

### Strengths and limitations

The strengths of this study are its large sample size, the good□quality score of studies included in the analysis, and no evidence of publication bias among the included studies. Some of the study limitations include: (1) Our search was restricted to those published in English only, so language bias could have flawed our findings, (2) The analysis was dependent on single-centred studies, which may not accurately reflect the true prevalence of the concurrent infection on the African continent, (3) The sample sizes for six studies ^23, 28, 33, 35, 45, 48^ were relatively small, so the sampling error could lead to substantial bias in the meta-analysis, (4) Our analysis showed significant heterogeneity among the studies due to too many outcomes, and (5) Subgroup analysis was not performed which could affect the accuracy of the results presented.

## CONCLUSION

In general, our study found a high prevalence of concurrent malaria and dengue fever among febrile patients in (sub-Saharan) Africa with significant variation across regions. Women and children were more affected population groups by the coinfection than men and adults, respectively. *P. falciparum* and dengue virus serotype 2 and 3 were the predominant aetiologies of this coinfection in the region. Healthcare workers should bear in mind the possibility of dengue infection as one of the differential diagnoses for acute febrile illness as well as the possibility of co-existent malaria and dengue in endemic areas. Our study also showed significantly higher odds of malaria infection were documented due to acute dengue co-infection in contrast to malaria mono-infection. Furthermore, no notable odds of acute dengue infection were reported due to malaria co-infection as compared to dengue mono-infection. Besides, high-quality multi-centre prospective studies are required to verify the above conclusions and to gain more insights into malaria and dengue virus co-infection in the continent.

## Data Availability

All data produced in the present work are contained in the manuscript.

## Acknowledgement

Not applicable.

## Contributorship

TG designed the study, wrote the statistical analysis plan, monitored the review process, interpreted the data, cleaned and analysed the data, and wrote the draft manuscript. TG and JD assessed studies for inclusion. ZM and AH reviewed and commented the draft paper. All authors have approved the final version. TG is the guarantor and takes responsibility for the content of this article.

## Patient consent

Consent was not required when conducting a systematic review.

## Ethics approval

This study did not require ethical approval as the data used have been published previously, and hence are already in the public domain.

## Data sharing

Extracted data will be available upon request to the corresponding author.

